# MORTALITY DISPARITY BY SOCIOECONOMIC POSITION IN PEOPLE WITH AND WITHOUT DIABETES: OPEN COHORT STUDIES IN FOUR HIGH-INCOME COUNTRIES

**DOI:** 10.1101/2025.03.14.25323661

**Authors:** Jonne G ter Braake, Rimke C Vos, Eeva-Liisa Røssell, Dianna J Magliano, Sarah H Wild, the Scottish Diabetes Research Network epidemiology group, David Walsh, Jedidiah I Morton, Susanne Boel Graversen, Henrik Støvring, Tinne Laurberg

## Abstract

**Background:** There have been mixed findings on whether mortality is socially patterned among people with diabetes. We compared all-cause mortality trends by socioeconomic position (SEP) among people with and without diabetes for 2004-2021 in four high-income countries.

**Methods:** We conducted open cohort studies in Australia, Denmark, the Netherlands, and Scotland and included national or regional populations aged 35-69 years. We used the European standard population in 2013 to calculate age-standardised mortality rates (ASMRs) by calendar year, SEP quintile, diabetes status, and sex. SEP quintiles were defined using standardised disposable household income in Denmark and the Netherlands, and area-based indices in Australia and Scotland. We calculated the age-standardised slope index of inequality (SII) and age-adjusted relative index of inequality (RII) using Poisson regression as absolute and relative measures of socioeconomic inequality respectively across the study populations stratified by calendar year, diabetes status, and sex.

**Results:** 208,011 deaths occurred during 17 million person years (py) of follow-up among 35-69 year olds with diabetes, and 1.1 million deaths during 298 million py of follow-up among people without diabetes. ASMRs generally increased with increasing deprivation and varied between 1.3 (95% CI: 1.2-1.4) deaths per 1000 py to 29.4 (95% CI: 26.0-32.8) deaths per 1000 py. We found absolute and relative mortality inequality among those with and without diabetes, both generally increased during the follow-up period.

**Conclusion:** Disparities in mortality by SEP increased during follow-up in most countries. Strategies are needed to reduce excess mortality associated with low SEP and diabetes and related socioeconomic inequality.

## INTRODUCTION

Type 2 diabetes incidence and its complications are found to be socially patterned in high income countries, with greater risk of both among those of lower socioeconomic position (SEP) (1–3). As a consequence, differing prevalence of diabetes and its associated higher risk of mortality may contribute to inequalities in mortality by SEP in whole populations (4). Evidence on whether mortality is similarly socially patterned among those with an existing diagnosis of diabetes is mixed (5–10). Studies from Denmark, the UK, and the US have reported inverse associations between SEP and mortality among people with diabetes (5–7, 10); however, studies from Finland and Italy found no differences in mortality rates across SEP among females with diabetes, and only a slight increase in mortality rates in the most deprived group compared to the least deprived group for males (8, 9). The authors of the Finnish and Italian papers argued that routine follow-up care within equitable healthcare systems may mitigate SEP-related differences in mortality among people with diabetes (8, 9).

However, much of this evidence is from the late 20th century. In recent years, several high-income countries have reported widening SEP disparities in all-cause mortality in the general population (11, 12). Factors contributing to these growing inequalities include austerity measures introduced after the 2008 recession and behavioural responses to poverty, such as smoking and alcohol consumption (11, 12). Increasing diabetes prevalence may also contribute to the SEP disparity in mortality. Moreover, despite having equitable healthcare systems, differences in diabetes care utilization by SEP have been reported in both Denmark and the Netherlands (13, 14).

In this analysis, we therefore describe contemporary all-cause mortality trends by SEP among people with and without diabetes for 2004 to 2021 in four high-income countries – Australia, Denmark, the Netherlands, and Scotland.

## METHODS

### Study design and material

We conducted open cohort studies of adults aged 35-69 years old in four high-income countries: Australia, Denmark, the Netherlands, and Scotland. These countries were included based on convenience sampling. They have different healthcare systems and use different measures of socioeconomic inequality (Electronic Supplemental Material (ESM) Table 1). Summary characteristics of the country-specific data sources, duration of follow-up, and definition of diabetes (type 1 and type 2 combined) are provided in Supplemental Table 2. Briefly, the definition of diabetes was based on a primary or secondary care diagnosis and/or medication prescription or purchase. Counts of deaths and population sizes were obtained from national death registries and national population registers. The upper age limit was chosen to minimise the effect of retirement on individual level income.

**Table 1:** Characteristics of the included population in 2013 of each country, by socioeconomic quintiles, sex and diabetes status.

|  | Males |  |  |  |  |  |  | Females |  |  |  |  |  |  |
| --- | --- | --- | --- | --- | --- | --- | --- | --- | --- | --- | --- | --- | --- | --- |
|  | No diabetes |  |  | Diabetes |  |  | Age-standardised diabetes prevalence (%) | No diabetes |  |  | Diabetes |  |  | Age-standardised diabetes prevalence (%) |
|  | Deaths | Person years | Mean age (SD) | Deaths | Person years | Mean age (SD) |  | Deaths | Person years | Mean age (SD) | Deaths | Person years | Mean age (SD) |  |
| Australia |  |  |  |  |  |  |  |  |  |  |  |  |  |  |
| Q1 (Most deprived) | 4330 | 768507 | 49.4 (11.6) | 911 | 65992 | 56.9 (10.4) | 5.5 | 2517 | 777898 | 49.4 (11.6) | 520 | 58865 | 55.3 (11.0) | 5.4 |
| Q2 | 3872 | 762197 | 49.6 (11.7) | 723 | 55558 | 56.8 (10.5) | 4.8 | 2252 | 770248 | 49.6 (11.6) | 370 | 46802 | 55.2 (11.1) | 4.4 |
| Q3 | 3079 | 747637 | 49.1 (11.6) | 620 | 54243 | 56.6 (10.5) | 4.7 | 1870 | 770604 | 49.0 (11.6) | 307 | 44146 | 55.1 (11.1) | 4.1 |
| Q4 | 2518 | 739106 | 48.3 (11.5) | 488 | 46335 | 56.6 (10.5) | 4.0 | 1628 | 756251 | 48.4 (11.5) | 232 | 36557 | 55.0 (11.1) | 3.4 |
| Q5 (Least deprived) | 2419 | 865438 | 48.4 (11.5) | 359 | 37120 | 56.7 (10.5) | 2.7 | 1636 | 903839 | 48.4 (11.5) | 145 | 28142 | 55.1 (11.2) | 2.2 |
| Denmark |  |  |  |  |  |  |  |  |  |  |  |  |  |  |
| Q1 (Most deprived) | 842 | 101287 | 46.2 (11.1) | 164 | 8605 | 53.9 (11.6) | 5.6 | 365 | 89446 | 46.4 (11.6) | 77 | 6070 | 53.9 (12.1) | 4.6 |
| Q2 | 1901 | 135931 | 50.2 (12.5) | 450 | 16776 | 58.2 (10.8) | 7.6 | 1197 | 167362 | 51.1 (13.0) | 206 | 14551 | 58.1 (11.1) | 5.7 |
| Q3 | 1616 | 228712 | 48.5 (12.2) | 450 | 20180 | 57.0 (11.0) | 5.4 | 1125 | 251594 | 49.4 (12.4) | 220 | 15363 | 56.3 (11.2) | 4.1 |
| Q4 | 1113 | 327850 | 47.9 (11.6) | 277 | 19591 | 56.2 (10.6) | 3.8 | 856 | 326884 | 48.4 (11.6) | 139 | 13132 | 54.4 (11.0) | 2.9 |
| Q5 (Least deprived) | 1092 | 415367 | 50.9 (11.3) | 214 | 20652 | 57.6 (9.3) | 3.3 | 780 | 401999 | 50.7 (11.0) | 60 | 11323 | 55.2 (10.2) | 2.2 |
| Netherlands |  |  |  |  |  |  |  |  |  |  |  |  |  |  |
| Q1 (Most deprived) | 2850 | 302478 | 48.4 (11.3) | 679 | 30438 | 56.0 (10.5) | 6.4 | 2105 | 321765 | 51.5 (11.7) | 458 | 32197 | 58.6 (9.5) | 6.4 |
| Q2 | 2914 | 406279 | 50.6 (12.3) | 772 | 45982 | 59.0 (10.1) | 6.8 | 2317 | 497545 | 51.5 (12.3) | 517 | 44536 | 58.7 (10.2) | 5.8 |
| Q3 | 3514 | 691770 | 50.6 (12.3) | 800 | 59262 | 58.6 (10.2) | 5.3 | 2598 | 732598 | 51.4 (12.2) | 416 | 47851 | 57.8 (10.6) | 4.5 |
| Q4 | 2985 | 985817 | 48.1<br>(11.6) | 561 | 57474 | 56.5<br>(10.6) | 3.7 | 2202 | 946066 | 48.1<br>(11.5) | 262 | 38866 | 54.9<br>(11.1) | 2.9 |
| Q5 (Least<br>deprived) | 2934 | 1240308 | 48.9<br>(10.9) | 425 | 55906 | 56.5<br>(9.7) | 2.9 | 1976 | 1205541 | 47.6<br>(10.6) | 166 | 31306 | 54.3<br>(10.4) | 1.8 |
| Scotland |  |  |  |  |  |  |  |  |  |  |  |  |  |  |
| Q1 (Most<br>deprived) | 2105 | 195423 | 48.4<br>(11.4) | 406 | 22465 | 54.8<br>(10.9) | 7.7 | 1325 | 215088 | 48.6<br>(11.5) | 256 | 17135 | 54.8<br>(11.1) | 5.6 |
| Q2 | 1554 | 206017 | 49.0<br>(11.6) | 349 | 21222 | 55.8<br>(10.8) | 6.8 | 1074 | 226140 | 49.3<br>(11.6) | 187 | 15429 | 55.8<br>(10.9) | 4.7 |
| Q3 | 1315 | 223660 | 49.4<br>(11.6) | 208 | 19358 | 56.4<br>(10.7) | 5.7 | 912 | 241177 | 49.7<br>(11.7) | 151 | 12736 | 55.9<br>(10.9) | 3.7 |
| Q4 | 1042 | 235402 | 49.3<br>(11.6) | 179 | 17355 | 56.6<br>(10.5) | 4.8 | 773 | 251921 | 49.5<br>(11.6) | 92 | 10672 | 56.1<br>(10.8) | 3.0 |
| Q5 (Least<br>deprived) | 766 | 238488 | 49.1<br>(11.6) | 114 | 14076 | 56.8<br>(10.5) | 3.9 | 646 | 257608 | 49.3<br>(11.6) | 59 | 8380 | 56.5<br>(10.9) | 2.3 |

### Data coverage and quality assessment

Australian data covered roughly 80% of the national population, as data were limited to those living in Victoria, New South Wales, the Australian Capital Territory, and Queensland.

Denmark, the Netherlands and Scotland provided national cohorts. The proportion with diabetes captured ranged from around 75% in the Netherlands, to >99% in Scotland (15–18).

### Procedures

Each country obtained routine data from their populations based on the following outline (for country-specific details, see Table 2): individuals were included at 1) the start of follow-up (2004-2008, varying by country, based on data availability); 2) the date they reached the age of 35 years or 3) migrated into the country, whichever occurred last. Individuals were followed until 1) date of death, 2) emigration, 3) reaching the age of 70 years, or 4) end of follow-up on 31 Dec 2021, whichever came first.

For each country, calendar year, sex (sex assigned at birth except for the Netherlands where last registered sex was used), age group (35-49, 50-54,…,65-69), diabetes status (yes/no) and quintile of SEP, we assessed time at risk and deaths during follow-up. People that developed diabetes during follow-up contributed person time to the appropriate groups before and after their date of diabetes diagnosis. Individual contributions of risk time and deaths were aggregated within the aforementioned categories and for cross-country comparison. Due to data safety and privacy concerns, categories with few events (n<5-10, depending on the country, ESM Table 2) were replaced by a random integer within the range.

Individual-level data were processed on designated servers provided by official authorities (Statistics Denmark, Statistics Netherlands, the Scottish diabetes research data safe haven, and the Secure Unified Research Environment for Australian data) and the institution responsible provided aggregated data for the country level comparisons.

### Definition of socioeconomic position

An individual-based approach (Denmark and the Netherlands) and an area-based approach (Australia and Scotland) were used in the definition of SEP. These two approaches were used because of data availability. The individual-level SEP measure used in Denmark and the Netherlands was based on standardised disposable household income, meaning the post-tax net amount a household can spend per year, adjusted for household size and composition. This income was assigned to all persons that were part of the household in the given year. We updated the individual-level SEP each year based on information of the previous year, thus individuals in the Danish and Dutch populations could change SEP strata during follow-up.

As area-based SEP measures, the Australian Index of Relative Socio-economic Disadvantage (IRSD) (19) and the Scottish Index of Multiple Deprivation (SIMD) (20) were used. These area-based deprivation measures consist of information across several domains, including (a selection of) income, employment, education, marital status, health, access to services, crime, housing, and access to internet (19, 20). One measure per person was used over the follow-up period, derived from a person’s most recent address.

All SEP measures were converted into quintiles. Individuals with missing SEP were excluded from analyses.

### Statistical analyses

We summarised country-specific basic demographics and the age-standardised diabetes prevalence by SEP in 2013 to provide an overview of the four populations. We calculated age-standardised mortality rates (ASMRs) for each calendar year by diabetes status, sex and SEP quintiles using direct standardisation and the 2013 European population (21) as the reference. We calculated 95% CIs based on estimated standard errors for rates (Wald type CIs). Additionally, we calculated the age-standardised Slope Index of Inequality (SII) and Relative Index of Inequality (RII) using the methods described by Moreno-Betancur et al. (22). Briefly, the SII estimates the absolute difference in mortality rates across the entire socioeconomic spectrum. It is calculated by assuming a linear relationship between socioeconomic rank and mortality rates in a Poisson model with an identity link, allowing for an estimation of the mortality difference between individuals at the least deprived and most deprived ends of the socioeconomic scale while considering the distribution across the whole population. The RII estimates the relative excess risk (ratio) across the entire socioeconomic spectrum again by assuming a linear relationship between socioeconomic rank and mortality, but now in a Poisson model with a logarithmic link. We stratified analyses by diabetes status, sex and calendar year in both model types, and included age as a covariate when estimating the RII. A graphical explanation of the SII and RII is provided in the ESM (ESM Figure 1). To explore whether the SII and RII changed over time, additional Poisson regression models were developed. For annual changes in the SII and RII, an interaction term between the SII or RII and calendar year was added to the previously described models to calculate the SII and RII.

**Figure 1:**
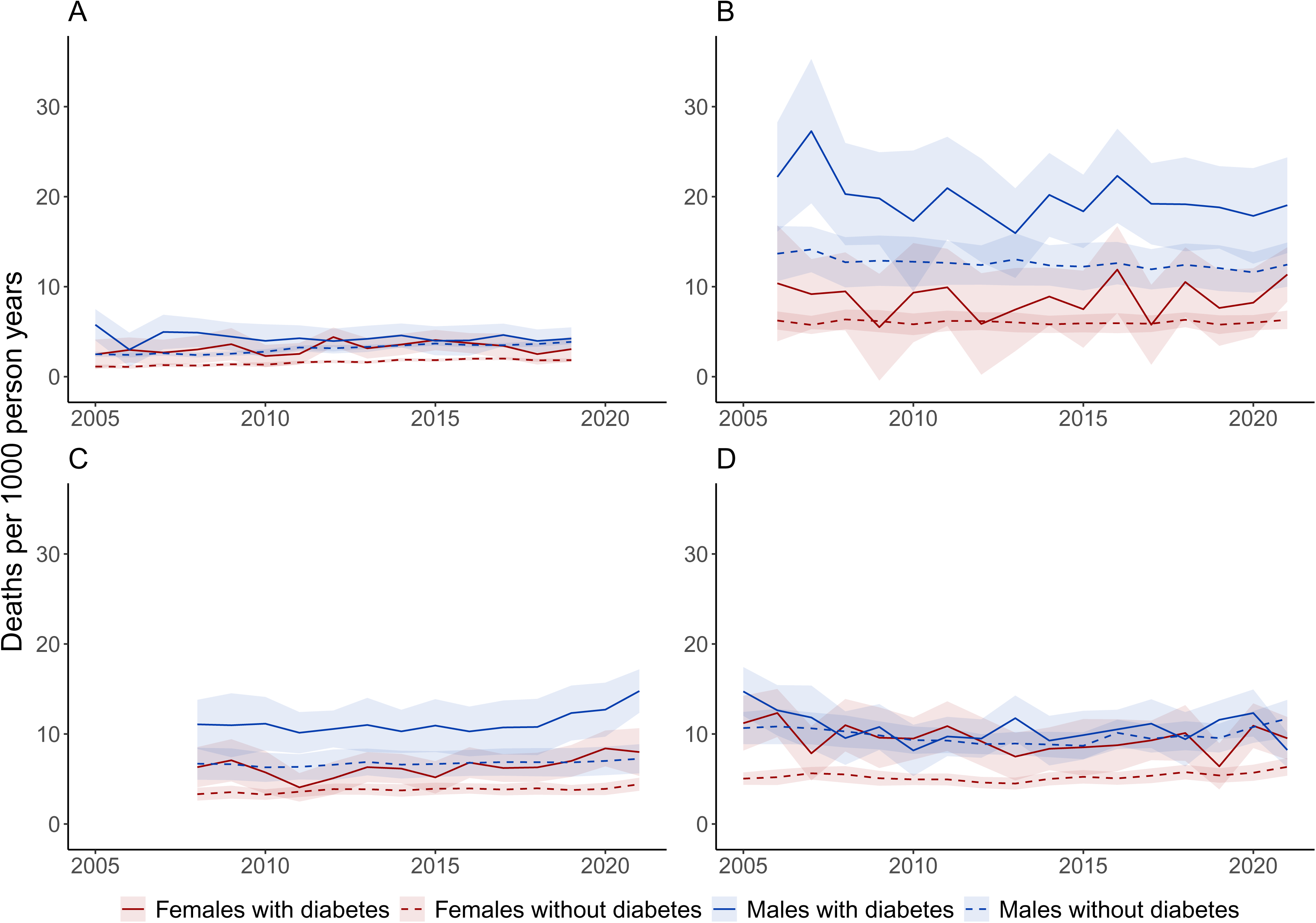
Age-standardised mortality rates by calendar year, for the least and most deprived quintiles by diabetes status, sex (left: males; right: females), and country, A: Australia, B: Denmark, C: Netherlands, and D: Scotland.

### Ethics approval

The Medical Ethical Review Board of Leiden University Medical Center mandated the review of research not falling under the Dutch Medical Research with Human Subjects Law (nWMO) to individual nWMO committees at Leiden University Medical Center. The nWMO committee reviewed the proposal and provided a declaration of no objection (non-WMO approval number: 24-3010). This study has been further approved by the scientific committee of the department of Public Health and Primary Care at Leiden University Medical Center.

### Data and Resource Availability

Aggregated data might be made available upon reasonable request to the corresponding author. There might be limitations on what the data can be used for, subject to approval from the data custodians. Researchers can request access to Scottish health records by contacting the electronic Data Research and Innovation Service (eDRIS). Details of the available datasets and the application process are available from

https://publichealthscotland.scot/services/data-research-and-innovation-services/electronic-data-research-and-innovation-service-edris. The code used for analyses are available via GitHub. We conducted all analyses in R-4.2.3 (23).

## RESULTS

After excluding person years with missing data on SEP (0.7% in Australia, 2.2% in Denmark, 2.1% in the Netherlands, and 0.3% in Scotland), we included 297,694,292 person years for people of 35-69 years of age without diabetes and 17,428,705 for people with diabetes for the four contributing countries for the period between 2004 and 2021. There were 1,136,842 and 208,011 deaths in people without and with diabetes, respectively. In all countries, the mean age of people with diabetes was higher compared to those without diabetes, males were slightly overrepresented in the group with diabetes and females in the group without diabetes, and there was a positive association between age-standardised diabetes prevalence and deprivation (as illustrated for 2013 in Table 1).

### Age-standardised mortality rates

In all countries and for both sexes, ASMRs generally increased with increasing deprivation and varied between 1.3 (95% CI: 1.2-1.4) deaths per 1000 person-years in the least deprived quintile of Danish females without diabetes in 2021 to 29.4 (95% CI: 26.0-32.8) deaths per 1000 person years in second to most deprived quintile of Danish males with diabetes in 2007 (Figure 1, ESM Table 3, and Figure 2). ASMRs were higher for people with compared to without diabetes and for males compared to females both for those with and without diabetes (Figure 1 and ESM Figure 2).

**Figure 2:**
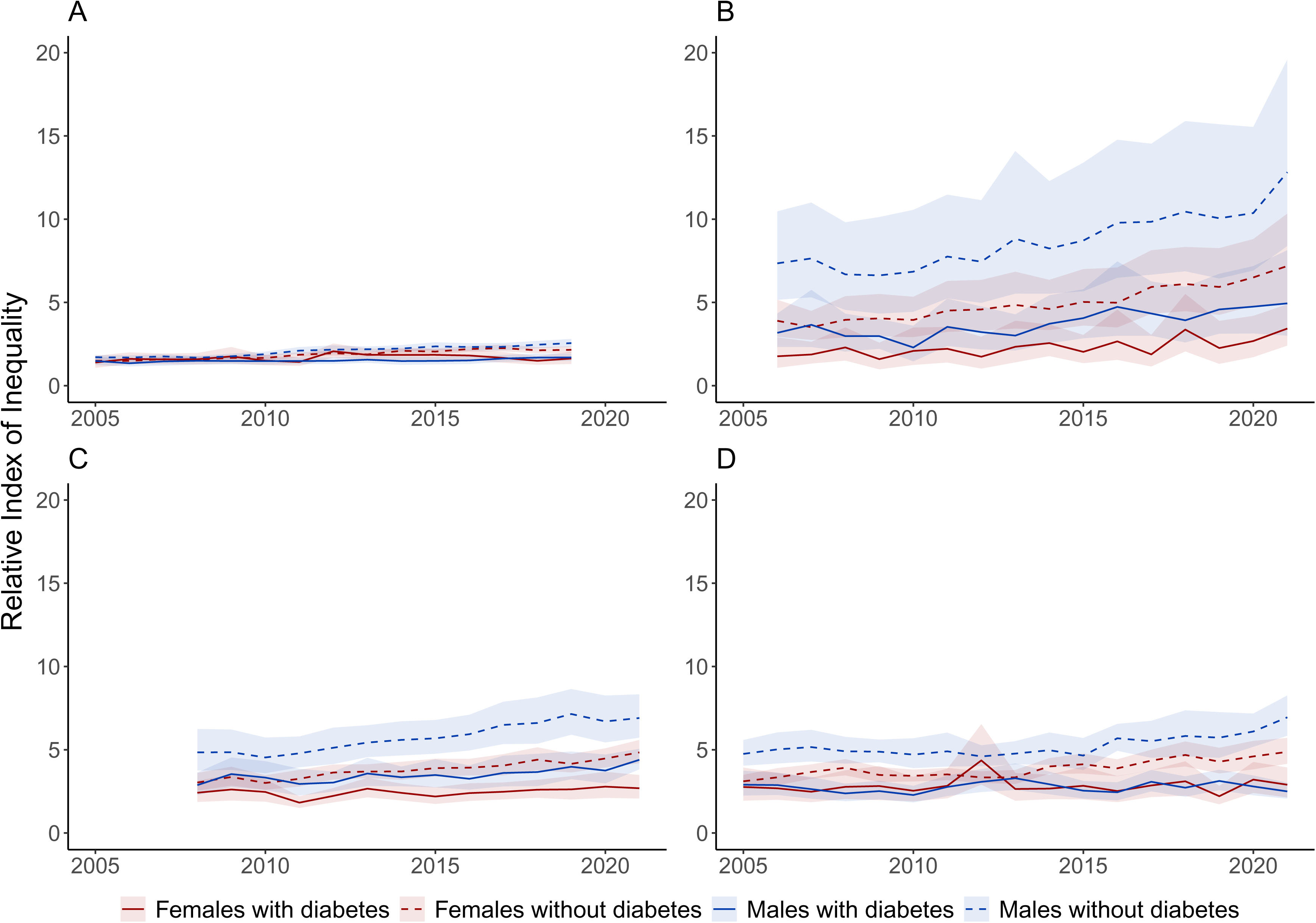
Slope index of inequality by diabetes status, sex, and country, A: Australia, B: Denmark, C: Netherlands, and D: Scotland.

### Slope index of inequality and relative index of inequality

Absolute mortality inequality, measured by the SII, was generally larger among people with compared to without diabetes, although 95% CIs generally overlapped (Figure 2). In people with diabetes, the SII ranged between 2.3 (95% CI: 1.1-3.5) deaths per 1000 person years (Australian females in 2010) and 27.3 (95% CI: 19.3-35.4) deaths per 1000 person years (Danish males in 2007) (ESM Table 4). Among people without diabetes the SII ranged from 1.1 (95% CI: 0.9-1.3) deaths per 1000 person years (Australian females in 2005) to 14.1 (95% CI: 11.6-16.7) deaths per 1000 person years (Danish males in 2007) (ESM Table 4). The SII tended to be higher among males compared to females for both diabetes statuses, with overlapping 95% CIs. During follow-up, we observed a general increase in SII per year, except for Australian females with diabetes and Scottish males with diabetes, where the SII remained stable (ESM Table 5). However, the annual increase in SII was smaller than 0.1 deaths per 1000 person years for all groups.

The RII was generally lower among people with diabetes compared to those without diabetes. The RII ranged between 1.35 (95% CI: 1.15-1.57 for Australian males in 2006) and 4.92 (95% CI: 3.00-8.06 for Danish males in 2021) in people with diabetes and between 1.49 (95% CI: 1.39-1.60 for Australian females in 2006) and 12.82 (95% CI: 8.39-19.59 for Danish males in 2021) in those without diabetes (Figure 3, ESM Table 6). Among people with diabetes, trends in RII were similar by sex in Australia and Scotland and higher among males compared to females in Denmark and the Netherlands, although 95% confidence intervals overlapped in most years. For people without diabetes, we observed a higher RII in males compared to females in all four countries, although 95% confidence intervals again overlapped in Australia and Denmark. During follow-up, we observed a general increase in the RII, except for Dutch females with diabetes and Australians and Scots of both sexes with diabetes, where the RII remained stable (ESM table 7). The annual change in the RII ranged from 1.6% (95% CI: 0.03-2.9% for Scottish males without diabetes) to 4.9% (95% CI: 3.2-6.7% for Danish females without diabetes) (ESM Table 7).

**Figure 3:**
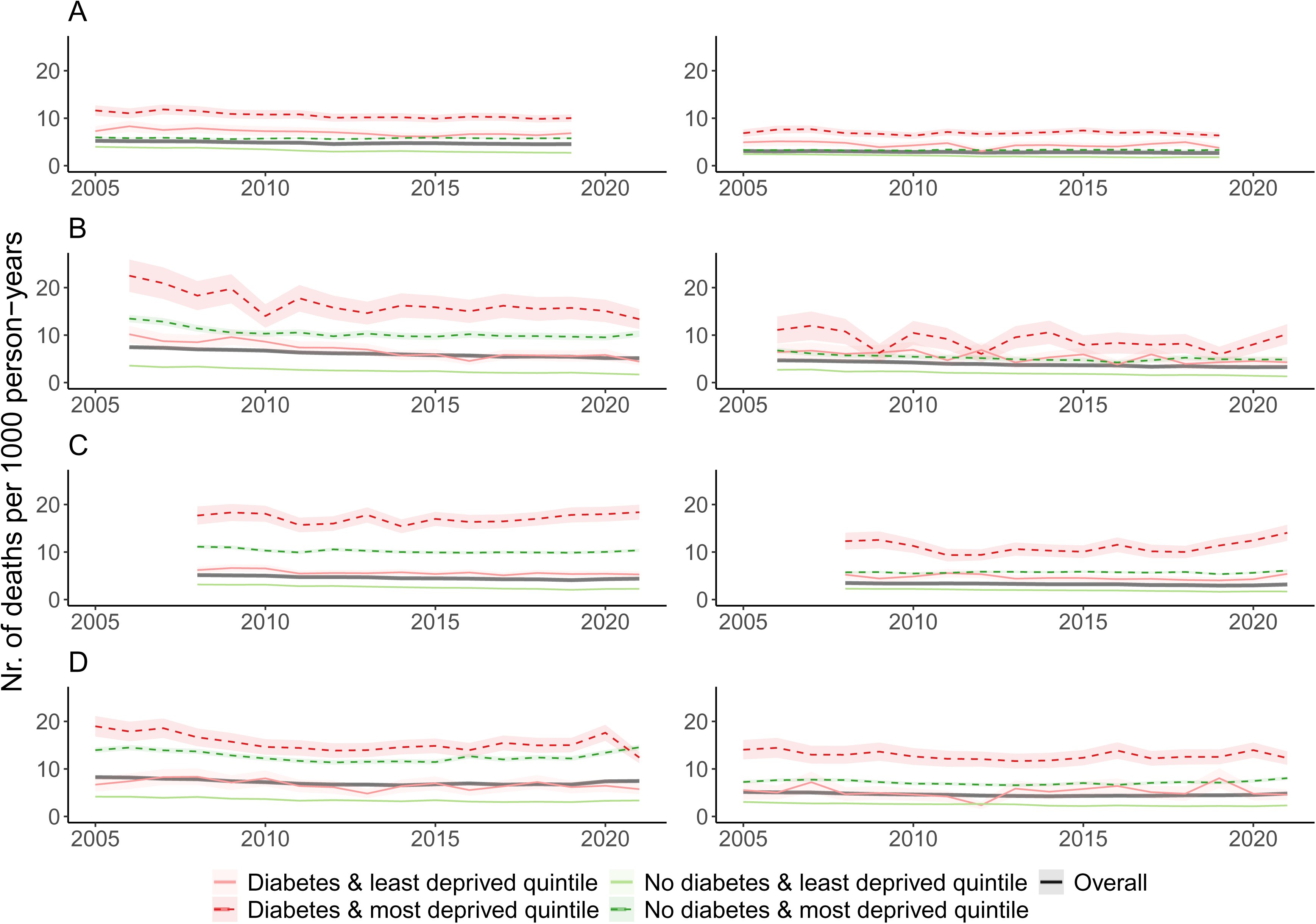
Relative index of inequality by diabetes status, sex, and country, A: Australia, B: Denmark, C: Netherlands, and D: Scotland.

## DISCUSSION

We compared all-cause mortality trends by SEP among people with and without diabetes in four high-income countries. Across countries, we observed increasing ASMRs with increasing deprivation, and higher ASMRs for people with compared to without diabetes and for males compared to females for those with and without diabetes. We found both absolute and relative mortality inequality by SEP irrespective of diabetes status with larger inequalities among males than females in some groups. Among people without diabetes, individuals at the most deprived end of the socioeconomic scale were estimated to have a 1.5 to almost 13 times higher mortality rate in the period of analysis compared to individuals at the least deprived end of the socioeconomic scale. For those with diabetes, this ranged between 1.4 and almost 5 times higher mortality rates. We generally observed an increase in absolute and relative mortality inequality over time. Although the annual increase in the measure of absolute mortality inequality remained small across all groups, we found a considerable annual increase in the measure of relative mortality inequality, ranging between an 8% to 27% increase over a 5-year period.

Relative mortality inequality was more pronounced among people without diabetes, whereas absolute mortality inequality was greater among people with diabetes. This can be explained by the differing mortality rates between those with versus without diabetes. Relative inequality is measured as a ratio, indicating that if the estimated mortality rates at both extremes of the socioeconomic scale are doubled, the relative risk remains unchanged, regardless of the actual number of deaths. In contrast, absolute inequality is measured as the difference in mortality rates and is therefore higher in groups where mortality is higher for a given relative risk and vice versa. The higher overall mortality rates in people with diabetes compared to those without diabetes contributes to the different patterns in absolute and relative measures of socioeconomic inequality we observed.

SEP-related mortality disparities persist after diabetes development, suggesting that the current health care received by those with diabetes does not sufficiently mitigate SEP-related differences in mortality. A more targeted approach may be needed to reduce SEP-related differences in mortality, including increased disease surveillance combined with social care and promoting equitable standard care utilization regardless of SEP (13, 14). This is especially urgent because we observed that SEP-related mortality differences are widening. In the four included countries, clinical guidelines do not consider treatment differentiation based on SEP although the ASSIGN cardiovascular risk score used in Scotland does include the Scottish measure of area-based deprivation (24–28). Therefore, strategies to effectively mitigate SEP-related differences in mortality among people with and without diabetes are required. Efforts to reduce inequalities across whole populations are also desirable based on our findings.

Although patterns of socioeconomic inequality are similar in the four included countries, the magnitude differs by country and in comparison to the findings of previous studies (5, 6, 10). Part of the variation can be explained by the use of different estimates for SEP. We used an individual-level measure in Denmark and the Netherlands, and area-based measures of SEP in Australia and Scotland. Comparability of these approaches is limited as previous research indicated that individuals may be classified differently depending on whether an area-based or individual-level measure is used (29). Specifically, McCartney et al. (2023) reported that in Scotland the majority of deprived citizens do not live in neighbourhoods classified as the most deprived (29). Further variation has been reported when comparing different individual-level measures of SEP, for example income, education, or occupational class (30). Therefore, generalisability of the magnitude of inequality might be limited, but the direction of SEP-related mortality inequality is consistent across contexts and measures.

A major strength of our study is the inclusion of data from whole populations in three countries and 80% of the population of a fourth country. Reliable ranking of SEP could be performed as established area-based proxies for SEP were used in Australia and Scotland, and the individual-level SEP in Denmark and the Netherlands was based on tax information, ensuring nearly complete coverage of the populations and quality of the data.

As mentioned above, the use of both individual-level and area-based measures for SEP in this study could be considered a limitation. We were limited by data availability and used the best measures available in each country. Another limitation is that we could not distinguish between type 1 and type 2 diabetes in all countries, and therefore grouped all diabetes together. As approximately 90% of diabetes is type 2 diabetes, the patterns we observed mostly pertain to type 2 diabetes. Additionally, in the Netherlands we only had data on medication prescriptions available to define diabetes diagnosis. Therefore, we estimate that we have missed around 20-30% of people with diabetes who are receiving lifestyle interventions only (16), who are likely to have lower mortality than people with diabetes who have been prescribed glucose-lowering drugs.

In conclusion, our study identifies marked SEP-related disparities in all-cause mortality among individuals with and without diabetes that appear to be widening over time. Further research is required to identify the role of other factors such as smoking status that may contribute to effect modification of the association between SEP and mortality by sex in some groups. To improve population health, strategies are needed to reduce excess mortality associated with socioeconomic disadvantage for people with and without diabetes.

## Supporting information

ESM

## Data Availability

Aggregated data might be made available upon reasonable request to the corresponding author. There might be limitations on what the data can be used for, subject to approval from the data custodians. Researchers can request access to Scottish health records by contacting the electronic Data Research and Innovation Service (eDRIS). Details of the available datasets and the application process are available from https://publichealthscotland.scot/services/data-research-and-innovation-services/electronic-data-research-and-innovation-service-edris.

https://github.com/Jonneterbraake/Diabetes-health-equity-project

## Abbreviations

ASMR: Age-standardised mortality rates
eDRIS: electronic Data Research and Innovation Service
ESM: Electronic Supplemental Material
IRSD: Australian Index of Relative Socio-economic Disadvantage
nWMO: Dutch Medical Research with Human Subjects Law
SEP: Socioeconomic position
SIMD: Scottish Index of Multiple Deprivation
SII: Slope Index of Inequality
RII: Relative Index of Inequality

## ACKNOWLEDGEMENTS

Scottish Care Information – diabetes [SCI-diabetes] data are available for service planning, audit and research thanks to people with diabetes who contribute their information, to the work of numerous NHS staff who enter the data and to members of key organisations (the SCI-Diabetes Steering Group, the Scottish Diabetes Group, diabetes managed clinical networks in each Health Board) involved in setting up, maintaining and overseeing SCI-diabetes system. The authors acknowledge the support of the electronic Data Research and Innovation Service within Public Health Scotland in the provisioning and linking of other data to SCI-diabetes that is funded by grants to Professor Helen Colhoun. The Scottish Diabetes Research Network receives financial support from NHS Research Scotland (NRS). We thank Philippa Haxton at National Records of Scotland for providing aggregated population data by age, sex, calendar year, and Scottish Index of Multiple Deprivation quintile. We acknowledge the work of Diabetes Australia and the Australian Institute of Health and Welfare for supplying the Australian data. This work was also partly supported by the Victoria State Government Operational Infrastructure Support Program. We would like to acknowledge the Danish Health Data Authority for providing the Danish data. We are grateful to Statistics Netherlands for providing and collecting the Dutch data. Results are based on calculations by the Leiden University Medical Center using non-public microdata from Statistics Netherlands.

## Role of study sponsor or funder

The study sponsor/funder was not involved in the design of the study; the collection, analysis, and interpretation of data; writing the report; and did not impose any restrictions regarding the publication of the report.

## Authors’ relationships and activities

Outside of the submitted work, HS received personal fees for statistical consulting from Novartis, Pfizer, MSD (Merck Sharpe & Dohme), Neumirna and Vesper Bio Aps. The remaining authors declare no competing interests.

## Contribution statement

HS, RCV, SHW, TL, and SBG conceived the study. HS, ER, and JGB coordinated data collection from the collaborating institutes. HS designed data analyses and HS and JGB conducted analyses. JGB drafted the manuscript. All authors contributed to the study design, data interpretation and editing of the report, and approved the final submitted version of the manuscript. TL had final responsibility for the decision to submit for publication. HS is the guarantor of this work and, as such, had full access to all the data in the study and takes responsibility for the integrity of the data and the accuracy of the data analysis.

