## Supplementary material for "MORTALITY DISPARITY BY SOCIOECONOMIC POSITION IN PEOPLE WITH AND WITHOUT DIABETES: OPEN COHORT STUDIES IN FOUR HIGH-INCOME COUNTRIES": ESM

**Electronic supplementary material (ESM)**

### ESM Table 1: Summary of healthcare systems and social characteristics in Australia, Denmark, the Netherlands and the United Kingdom.

|  | **OECD classification of the healthcare system (1)** | **Life expectancy at birth (2)** | **Poverty rate^-^ (3)** | **Smoking (% of population aged 15 and over) (2)** | **Alcohol consumption^+^ (2)** | **Overweight (% among adults) (2)** |
| --- | --- | --- | --- | --- | --- | --- |
| **Australia** | National Health Insurance | 83 | 0.126 | 11.2 | 9.5 | 65.2 |
| **Denmark** | National Health Service | 81.5 | 0.065 | 16.9 | 9.5 | 48.8 |
| **Netherlands** | Etatist Social Health Insurance | 82.2 | 0.085 | 15.4 | 8.2 | 48.4 |
| **United Kingdom** | National Health Service | 81.4 | 0.117 | 15.8 | 9.7 | 64.2 |

^-^ The ratio of the number of people (in a given age group) whose income falls below the poverty line; taken as half the median household income of the total population

^+^ Annual sales of pure alcohol in liters per person aged 15 years and over

### ESM Table 2: Summary of the characteristics of the data of the included countries

| **Country** | **Origin of data** | **Years included in analysis** | **Definition of diabetes** | **Level of combining deaths** | **Inclusion of migration data** |
| --- | --- | --- | --- | --- | --- |
| Australia | National Diabetes Service Scheme and population estimates | 2005-2019 | Clinical diagnosis^1^ certified by a doctor, nurse or credentialed diabetes educator | <5 in population without diabetes, <6 in population with diabetes | Not available |
| Denmark | Statistics Denmark | 2006-2021 | Register for Selected Chronic Diseases, an algorithm based on diabetes diagnoses^1^ and medication prescriptions (4) | <5 | Included |
| Netherlands | Statistics Netherlands | 2008-2021 | Receipt of an insulin or glucose-lowering drug prescription for at least 2 years | <10 | Included, registration to an address is used as a proxy |
| Scotland | SCI-Diabetes database and National Records of Scotland | 2004-2021 | Clinical diagnosis^1^ and primary care disease codes | <5 | Not available |

^1^ Diagnostic criteria for a diabetes diagnosis adhere to the World Health Organisation (WHO)/International Diabetes Federation (IDF) recommendations 2006 (5).

### ESM table 3: Extremes of age-standardised mortality rates (deaths per 1000 person-years) and 95% confidence intervals by year over the follow-up period.

|  | Males | | | | | | | | Females | | | | | | | |
| --- | --- | --- | --- | --- | --- | --- | --- | --- | --- | --- | --- | --- | --- | --- | --- | --- |
|  | No diabetes | | | | Diabetes | | | | No diabetes | | | | Diabetes | | | |
|  | Highest ASMR | Year | Lowest ASMR | Year | Highest ASMR | Year | Lowest ASMR | Year | Highest ASMR | Year | Lowest ASMR | Year | Highest ASMR | Year | Lowest ASMR | Year |
| Australia |  |  |  |  |  |  |  |  |  |  |  |  |  |  |  |  |
| Q1 (Most deprived) | 6.0 (5.8-6.20 | 2005 | 5.6 (5.4-5.8) | 2012 | 11.8 (10.8-12.9) | 2007 | 9.9 (9.1-10.6) | 2018 | 3.4 (3.3-3.5) | 2011 | 3.2 (3.0-3.3) | 2010 | 7.7 (6.9-8.5) | 2007 | 6.3 (5.7-7.0) | 2010 |
| Q2 | 5.5 (5.3-5.6) | 2005 | 4.9 (4.7-5.0) | 2012 | 11.6 (10.4-12.8) | 2005 | 8.8 (8.1-9.6) | 2018 | 3.2 (3.1-3.4) | 2005 | 2.9 (2.8-3.0) | 2019 | 7.8 (6.9-8.7) | 2008 | 6.0 (5.3-6.7) | 2010 |
| Q3 | 4.8 (4.6-5.0) | 2005 | 4.0 (3.8-4.1) | 2012 | 10.8 (9.7-12.0) | 2006 | 7.6 (6.8-8.4) | 2012 | 2.9 (2.8-3.0) | 2005 | 2.3 (2.2-2.4) | 2019 | 6.3 (5.5-7.2) | 2008 | 5.3 (4.7-6.0) | 2013 |
| Q4 | 4.4 (4.3-4.6) | 2006 | 3.5 (3.3-3.6) | 2019 | 9.8 (8.7-10.9) | 2007 | 7.1 (6.4-7.9) | 2019 | 2.7 (2.6-2.9) | 2005 | 2.1 (2.0-2.2) | 2017 | 6.4 (5.4-7.3) | 2007 | 4.8 (4.1-5.5) | 2012 |
| Q5 (Least deprived) | 4.0 (3.8-4.1) | 2005 | 2.7 (2.6-2.8) | 2019 | 8.3 (7.1-9.5) | 2006 | 6.2 (5.4-6.9) | 2015 | 2.4 (2.3-2.6) | 2005 | 1.7 (1.6-1.8) | 2017 | 5.1 (4.2-6.1) | 2006 | 3.1 (2.4-3.7) | 2012 |
| Denmark |  |  |  |  |  |  |  |  |  |  |  |  |  |  |  |  |
| Q1 (Most deprived) | 13.5 (12.7-14.3) | 2006 | 9.5 (8.9-10.2) | 2020 | 22.5 (19.1-25.9) | 2006 | 13.4 (11.3-15.4) | 2021 | 6.8 (6.2-7.3) | 2006 | 4.2 (3.8-4.7) | 2016 | 12.0 (9.0-15.0) | 2007 | 6.1 (4.3-7.9) | 2012 |
| Q2 | 15.1 (14.5-15.8) | 2006 | 11.5 (11.0-12.1) | 2020 | 29.4 (26.0-32.8) | 2007 | 18.0 (15.8-20.1) | 2013 | 7.0 (6.6-7.5) | 2009 | 5.8 (5.4-6.2) | 2019 | 15.0 (12.1-17.0) | 2009 | 8.4 (6.9-9.9) | 2013 |
| Q3 | 8.3 (7.9-8.7) | 2009 | 5.8 (5.5-6.2) | 2021 | 24.4 (21.7-27.1) | 2007 | 12.6 (11.1-14.2) | 2021 | 5.2 (4.9-5.4) | 2006 | 3.8 (3.6-4.1) | 2021 | 16.6 (14.0-19.1) | 2006 | 8.9 (7.5-10.3) | 2018 |
| Q4 | 5.0 (4.7-5.3) | 2007 | 3.0 (2.8-3.2) | 2021 | 13.4 (11.6-15.1) | 2009 | 6.1 (5.1-7.1) | 2021 | 3.6 (3.4-3.8) | 2007 | 2.2 (2.0-2.4) | 2019 | 10.7 (8.7-12.8) | 2009 | 5.2 (4.1-6.4) | 2018 |
| Q5 (Least deprived) | 3.6 (3.4-3.8) | 2006 | 1.7 (1.6-1.8) | 2021 | 10.2 (8.4-11.9) | 2006 | 4.6 (3.8-5.4) | 2021 | 2.8 (2.6-2.9) | 2007 | 1.3-1.2-1.4) | 2021 | 7.2 (5.3-9.1) | 2006 | 4.0 (3.0-5.0) | 2016 |
| Netherlands |  |  |  |  |  |  |  |  |  |  |  |  |  |  |  |  |
| Q1 (Most deprived) | 11.1 (10.7-11.5) | 2008 | 9.9 (9.5-10.2) | 2016 | 18.4 (16.8-19.9) | 2021 | 15.4 (14.0-16.9) | 2014 | 6.1 (5.8-6.3) | 2021 | 5.3 (5.1-5.6) | 2019 | 14.0 (12.3-15.7) | 2021 | 9.3 (8.0-10.7) | 2011 |
| Q2 | 6.5 (6.2-6.7) | 2008 | 5.4 (5.2-5.7) | 2019 | 13.7 (12.3-15.1) | 2021 | 9.0 (8.1-10.0) | 2017 | 4.0 (3.8-4.2) | 2012 | 3.5 (3.4-3.7) | 2019 | 8.2 (7.2-9.2) | 2021 | 6.3 (5.6-7.1) | 2018 |
| Q3 | 4.7 (4.6-4.9) | 2008 | 3.6 (3.5-3.8) | 2019 | 9.3 (8.4-10.3) | 2021 | 7.4 (6.7-8.1) | 2016 | 3.2 (3.1-3.4) | 2011 | 2.7 (2.6-2.9) | 2019 | 7.0 (6.1-7.8) | 2018 | 5.6 (4.9-6.3) | 2013 |
| Q4 | 3.9 (3.7-4.0) | 2008 | 2.8 (2.7-2.9) | 2019 | 8.2 (7.4-9.0) | 2008 | 5.9 (5.3-6.5) | 2019 | 2.8 (2.7-2.9) | 2008 | 2.1 (2.0-2.1) | 2020 | 6.3 (5.5-7.0) | 2021 | 4.8 (4.2-5.5) | 2018 |
| Q5 (Least deprived) | 3.2 (3.0-3.3) | 2008 | 2.0 (2.0-2.1) | 2019 | 6.6 (5.9-7.4) | 2009 | 5.1 (4.5-5.7) | 2017 | 2.3 (2.2-2.4) | 2008 | 1.6 (1.5-1.7) | 2019 | 5.6 (4.7-6.4) | 2011 | 4.0 (3.3-4.7) | 2019 |
| Scotland |  |  |  |  |  |  |  |  |  |  |  |  |  |  |  |  |
| Q1 (Most deprived) | 14.6 (14.0-15.1) | 2021 | 11.4 (10.9-11.9) | 2012 | 19.8 (17.5-22.1) | 2004 | 12.4 (11.2-13.6) | 2021 | 8.1 (7.7-8.4) | 2021 | 6.6 (6.2-6.9) | 2013 | 14.8 (12.5-17.1) | 2004 | 11.7 (10.1-13.2) | 2013 |
| Q2 | 9.9 (9.4-10.3) | 2004 | 7.5 (7.1-7.9) | 2014 | 13.9 (12.1-15.8) | 2006 | 9.9 (8.6-11.2) | 2014 | 5.9 (5.5-6.2) | 2005 | 4.8 (4.5-5.1) | 2020 | 13.2 (11.1-15.4) | 2005 | 8.6 (7.4-9.7) | 2021 |
| Q3 | 7.4 (7.1-7.8) | 2004 | 5.5 (5.2-5.8) | 2014 | 13.4 (11.4-15.4) | 2005 | 7.9 (6.9-9.0) | 2021 | 4.5 (4.2-4.8) | 2004 | 3.5 (3.3-3.7) | 2014 | 10.8 (8.7-12.4) | 2006 | 5.9 (4.7-7.1) | 2012 |
| Q4 | 5.8 (5.5-6.2) | 2004 | 4.2 (4.0-4.5) | 2018 | 10.3 (8.7-11.9) | 2020 | 6.1 (4.9-7.2) | 2012 | 3.8 (3.5-4.1) | 2006 | 2.8 (2.6-3.0) | 2018 | 8.2 (6.4-10.1) | 2005 | 5.1 (4.0-6.3) | 2012 |
| Q5 (Least deprived) | 4.3 (4.0-4.6) | 2004 | 3.0 (2.8-3.2) | 2019 | 8.4 (6.6-10.1) | 2008 | 4.8 (3.9-5.8) | 2013 | 3.1 (2.8-3.3) | 2005 | 2.1 (2.0-2.3) | 2020 | 8.0 (5.9-10.1) | 2019 | 2.4 (1.6-3.2) | 2012 |

### ESM table 4: Range of the slope index of inequality (deaths per 1000 person-years) and 95% confidence intervals over the follow-up period, per country, diabetes status and sex.

|  | No diabetes | | | | Diabetes | | | |
| --- | --- | --- | --- | --- | --- | --- | --- | --- |
|  | Lowest SII | Year | Highest SII | Year | Lowest SII | Year | Highest SII | Year |
| Australia |  |  |  |  |  |  |  |  |
| Males | 2.40 (2.14-2.66) | 2008 | 3.86 (3.62-4.10) | 2019 | 3.01 (1.13-4.89) | 2006 | 5.76 (4.03-7.50) | 2005 |
| Females | 1.09 (0.88-1.29) | 2006 | 2.01 (1.84-2.17) | 2017 | 2.29 (1.06-3.53) | 2010 | 4.40 (3.34-5.46) | 2012 |
| Denmark |  |  |  |  |  |  |  |  |
| Males | 11.59 (9.33-13.84) | 2020 | 14.13 (11.61-16.66) | 2007 | 15.95 (10.97-20.92) | 2013 | 27.28 (19.27-35.30) | 2007 |
| Females | 5.75 (4.74-6.76) | 2007 | 6.36 (5.27-7.45) | 2008 | 5.34 (-0.52-11.19) | 2009 | 11.59 (6.75-16.42) | 2016 |
| Netherlands |  |  |  |  |  |  |  |  |
| Males | 6.30 (4.68-7.91) | 2010 | 7.25 (5.63-8.86) | 2021 | 10.14 (7.84-12.44) | 2011 | 14.77 (12.36-17.18) | 2021 |
| Females | 3.27 (2.68-3.85) | 2010 | 4.42 (3.69-5.14) | 2021 | 4.07 (2.49-5.65) | 2011 | 8.52 (6.54-10.50) | 2020 |
| Scotland |  |  |  |  |  |  |  |  |
| Males | 8.70 (7.21-10.18) | 2015 | 11.70 (9.61-13.79) | 2021 | 8.17 (5.32-11.02) | 2010 | 14.73 (12.02-17.43) | 2005 |
| Females | 4.49 (3.83-5.16) | 2013 | 6.34 (5.37-7.32) | 2021 | 6.41 (3.84-8.97) | 2019 | 12.33 (9.67-15.00) | 2006 |

### ESM table 5: Annual change in the slope index of inequality, per country, diabetes status and sex, including 95% confidence intervals.

| Country | Diabetes status | Sex | Change in slope index of inequality per year (deaths per 1000 person years) (95% CI) |
| --- | --- | --- | --- |
| Australia | No diabetes | Males | 7.8E-5 (6.2E-5−9.4E-5) |
|  |  | Females | 6.9E-5 (4.9E-5−8.9E-5) |
|  | Diabetes | Males | 1.1E-2 (1.0E-2−1.2E-2) |
|  |  | Females | 1.2E-3 (-3.5E-4−2.7E-3) |
| Denmark | No diabetes | Males | 8.9E-4 (8.1E-4−9.7E-4) |
|  |  | Females | 2.4E-3 (2.3E-3−2.5E-3) |
|  | Diabetes | Males | 5.5E-3 (2.1E-3−8.8E-3) |
|  |  | Females | 2.9E-2 (2.1E-2−3.9E-2) |
| Netherlands | No diabetes | Males | 2.4E-3 (2.3E-3−2.4E-3) |
|  |  | Females | 1.1E-3 (1.1E-3−1.2E-3) |
|  | Diabetes | Males | 9.6E-3 (8.3E-3−1.1E-2) |
|  |  | Females | 2.3E-3 (1.5E-4−4.4E-3) |
| Scotland | No diabetes | Males | 3.4E-3 (3.3E-3−3.5E-3) |
|  |  | Females | 3.3E-3 (3.2E-3−3.4E-3) |
|  | Diabetes | Males | -2.2E-4 (-3.5E-3−3.1E-3) |
|  |  | Females | 8.7E-3 (2.7E-3−1.5E-2) |

### ESM table 6: Extremes by calendar year of relative index of inequality and 95% confidence intervals over the follow-up period, by country, diabetes status and sex.

|  | No diabetes | | | | Diabetes | | | |
| --- | --- | --- | --- | --- | --- | --- | --- | --- |
|  | Lowest RII | Year | Highest RII | Year | Lowest RII | Year | Highest RII | Year |
| Australia |  |  |  |  |  |  |  |  |
| Males | 1.68 (1.58-1.79) | 2008 | 2.56 (2.33-2.81) | 2019 | 1.35 (1.15-1.57) | 2006 | 1.69 (1.49-1.92) | 2019 |
| Females | 1.49 (1.39-1.60) | 2006 | 2.25 (2.05-2.47) | 2017 | 1.39 (1.08-1.79) | 2005 | 2.07 (1.69-2.53) | 2012 |
| Denmark |  |  |  |  |  |  |  |  |
| Males | 6.62 (4.33-10.13) | 2009 | 12.82 (8.39-19.59) | 2021 | 2.29 (1.46-3.60) | 2010 | 4.92 (3.00-8.06) | 2021 |
| Females | 3.51 (2.75-4.49) | 2007 | 7.19 (5-10.33) | 2021 | 1.57 (0.98-2.53) | 2009 | 3.43 (2.41-4.89) | 2021 |
| Netherlands |  |  |  |  |  |  |  |  |
| Males | 4.54 (3.59-5.73) | 2010 | 7.15 (5.91-8.65) | 2019 | 2.88 (2.28-3.64) | 2008 | 4.40 (3.82-5.05) | 2021 |
| Females | 3.01 (2.6-3.47) | 2010 | 4.84 (4.19-5.59) | 2021 | 1.82 (1.51-2.20) | 2011 | 2.79 (2.11-3.70) | 2020 |
| Scotland |  |  |  |  |  |  |  |  |
| Males | 4.60 (4.02-5.27) | 2012 | 6.95 (5.84-8.26) | 2021 | 2.28 (1.84-2.83) | 2010 | 3.32 (2.59-4.25) | 2004 |
| Females | 3.11 (2.76-3.49) | 2005 | 4.87 (4.16-5.71) | 2021 | 2.21 (1.73-2.83) | 2019 | 4.36 (2.90-6.54) | 2012 |

### ESM table 7: Annual change in the relative index of inequality, per country, diabetes status and sex, including 95% confidence intervals.

| Country | Diabetes status | Sex | Change in relative index of inequality per year (%) |
| --- | --- | --- | --- |
| Australia | No diabetes | Males | 3.21 (2.10−4.32) |
|  |  | Females | 3.13 (2.18−4.10) |
|  | Diabetes | Males | 1.06 (-0.24−2.38) |
|  |  | Females | 0.65 (-0.81−2.14) |
| Denmark | No diabetes | Males | 4.04 (1.86−6.26) |
|  |  | Females | 4.92 (3.20−6.66) |
|  | Diabetes | Males | 3.75 (1.58−5.96) |
|  |  | Females | 3.50 (1.07−5.99) |
| Netherlands | No diabetes | Males | 3.60 (2.21−5.01) |
|  |  | Females | 4.11 (3.12−5.12) |
|  | Diabetes | Males | 2.29 (0.53−4.08) |
|  |  | Females | 1.53 (-0.22−3.32) |
| Scotland | No diabetes | Males | 1.58 (0.30−2.88) |
|  |  | Females | 2.23 (1.22−3.25) |
|  | Diabetes | Males | 0.00 (-1.57−1.35) |
|  |  | Females | 0.25 (-1.36−1.89) |

### ESM Figure 1: Graphical explanation of the slope index of inequality and relative index of inequality.

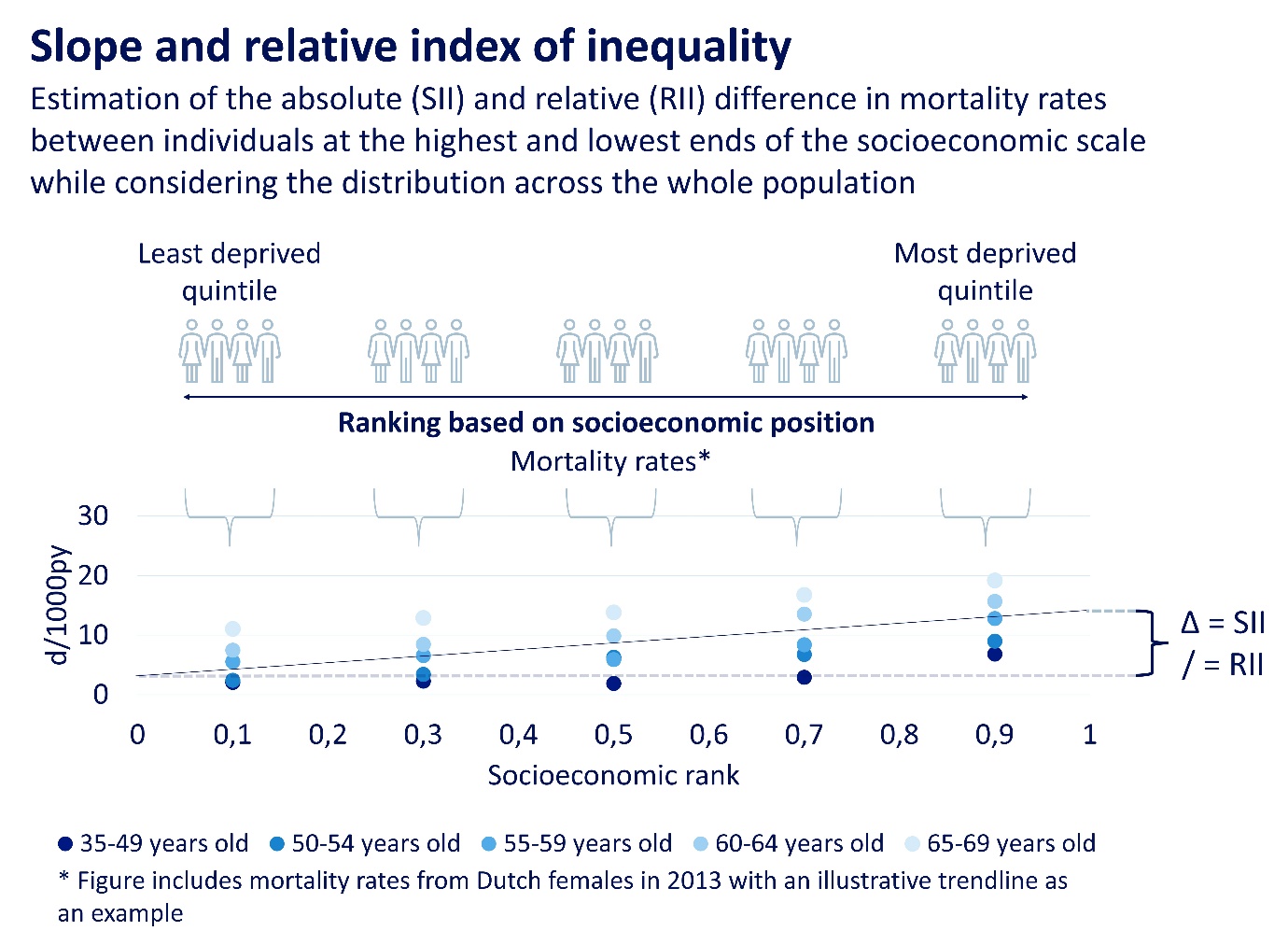

ESM figure 2: Age-standardised mortality rates by calendar year, for all five socioeconomic quintiles of those with and without diabetes, by country and sex.

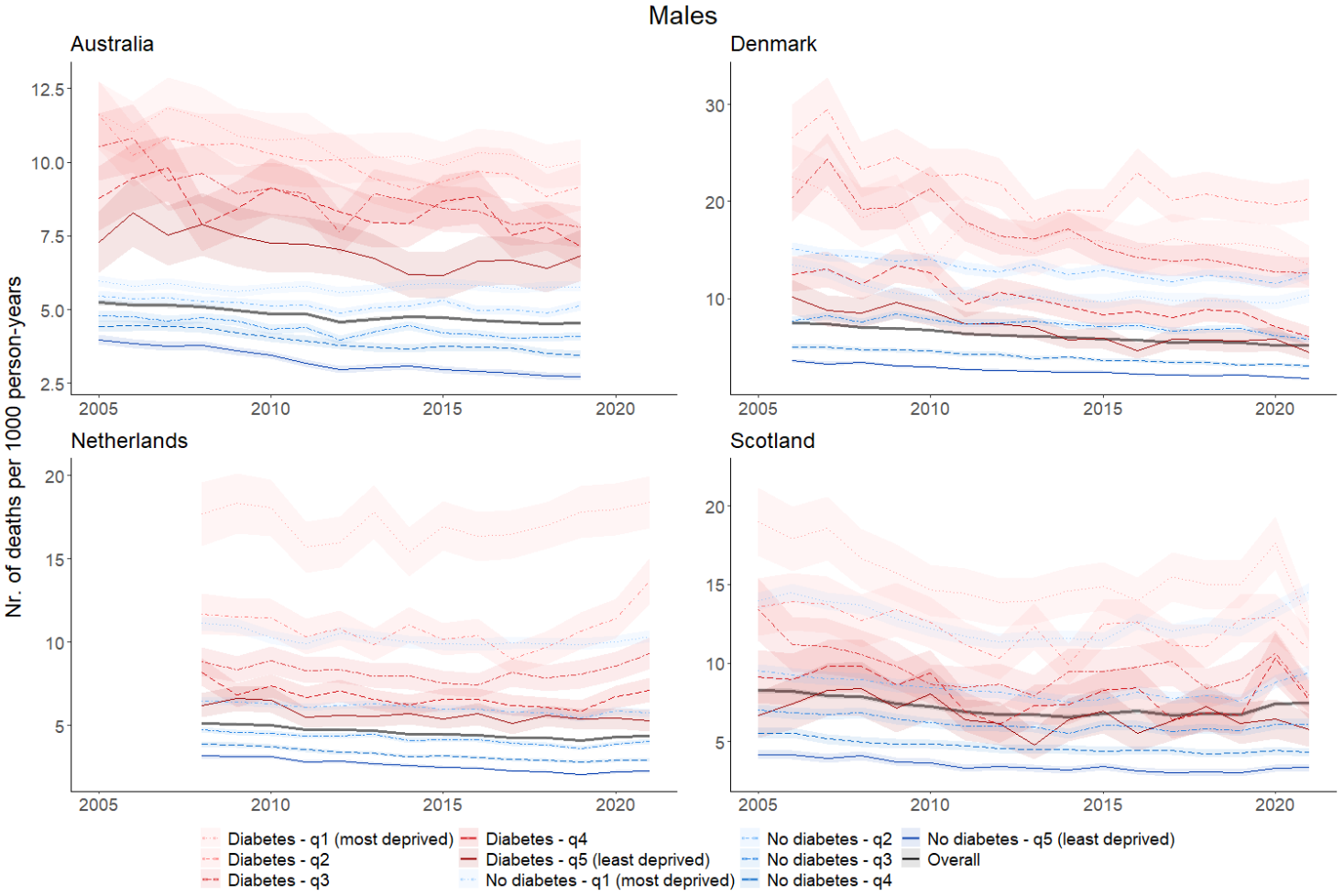

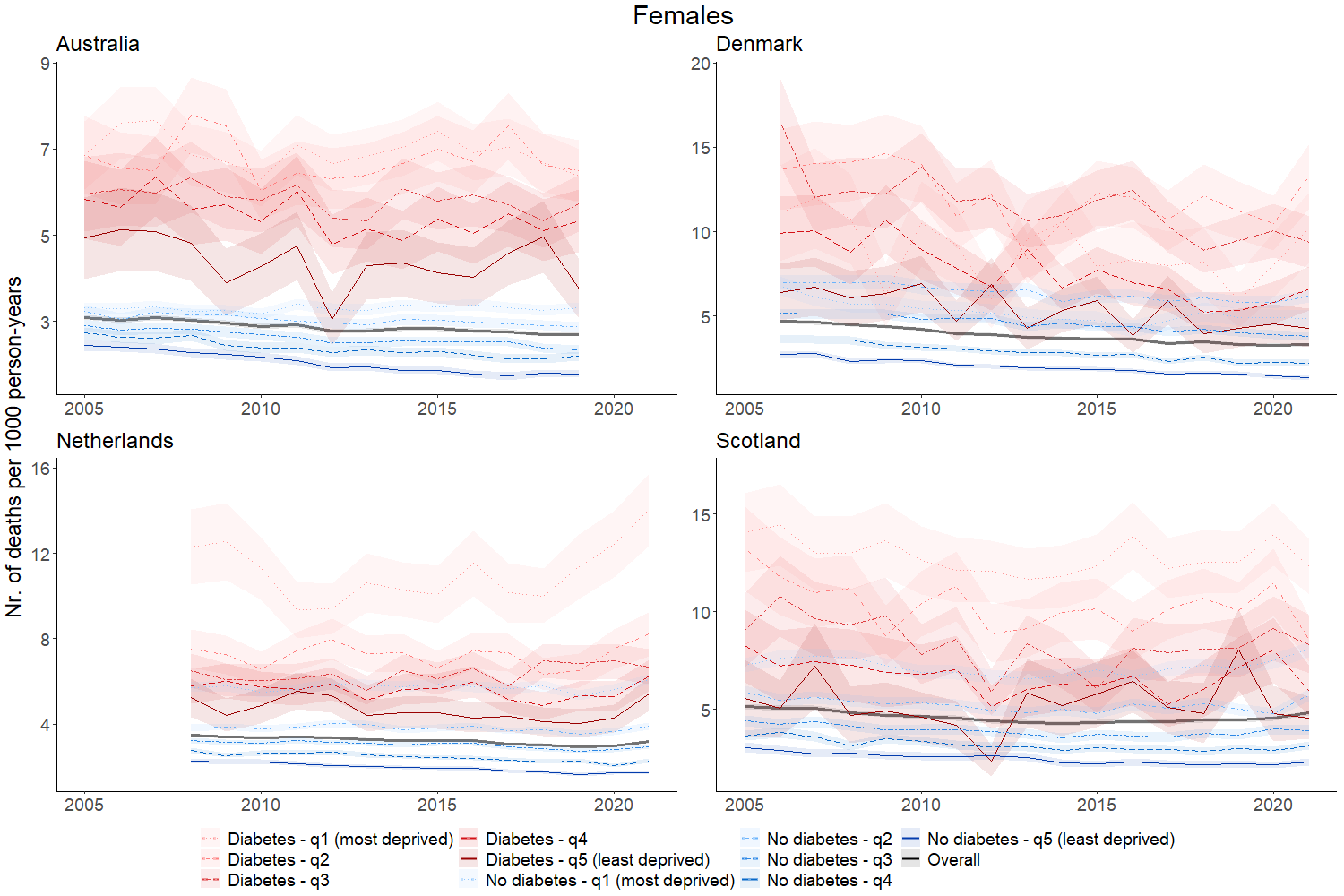

2.	OECD. Health at a Glance 2023: OECD Indicators. Paris: OECD Publishing; 2023.

3.	OECD. Poverty rate (indicator) [cited 2024 11 April]. Available from: <https://data.oecd.org/inequality/poverty-rate.htm#:~:text=The%20poverty%20rate%20is%20the,income%20of%20the%20total%20population>.

4.	Isaksen AA, Sandbæk A, Bjerg L. Validation of Register-Based Diabetes Classifiers in Danish Data. Clinical Epidemiology. 2023;Volume 15:569-81.

5.	Definition and diagnosis of diabetes mellitus and intermediate hyperglycemia. Repot of a WH/ID Consultation. 2006.
